# Comparison of the extent and progression of respiratory and cardiovascular disease in World Trade Center responders to lung screening participants

**DOI:** 10.1101/2024.10.25.24316091

**Authors:** Artit Jirapatnakul, Rowena Yip, Andrea Branch, David F Yankelevitz, Claudia I Henschke

## Abstract

Responders to the World Trade Center (WTC) site in the aftermath of the 9/11 attacks were exposed to toxic dust, which has been linked to increased risk of respiratory and cardiovascular disease. The respiratory and cardiovascular effects of WTC dust exposure have been studied using pulmonary function tests and the number of cardiovascular events, but computed tomography (CT) scans provide an opportunity to see the early structural changes in the lungs and cardiovascular system before clinical symptoms appear. CT scans are used in the screening and evaluation of respiratory diseases such as lung cancer, interstitial lung disease, and chronic obstructive pulmonary disease, and to visualize coronary arteries and quantify the amount of coronary artery calcifications; in fact, it is possible to detect multiple diseases from a single chest CT scan. While manual evaluation by a radiologist is often the gold standard, automated image analysis tools can quickly and accurately quantify these diseases.

We identified non-contrast chest CT scans from members of the World Trade Center General Responders Cohort (WTC GRC) with slice thickness of 2.5 mm or less. We used the open-source Chest Imaging Platform software to compute measures of emphysema and interstitial lung disease and research software from Cornell University to compute measures of pulmonary hypertension and coronary artery calcification. We identified a sex, age (within 5 years), smoking status, one or more CT scans, and follow-up time -matched cohort of participants enrolled in the lung screening program at Mount Sinai. We compared disease measures from the WTC GRC group to the lung screening group to assess whether there was a difference in the extent and progression of disease.

There were 4909 chest CT images of members of the WTC GRC that met our image quality criteria. There were 3855 members of the GRC for which we could obtain both chest CT images and clinical data. Of these, there were 2284 members for which we could obtain pulmonary disease measurements on at least one scan, 1246 members for which we could calculate cardiac measurements. The matched controls from the lung screening cohort consisted of 557 participants with 1122 chest CT images that met our image quality criteria and for which we obtained all four disease measures.

We compared members of the WTC GRC with matched participants from the lung screening program. One of the key findings is that after a median time of 11-13 years after 9/11, the WTC GRC group exhibited higher burdens of coronary artery calcification, emphysema, and interstitial lung disease compared with a matched control group of lung screening participants. This supports the continued surveillance of WTC responders.

## Introduction

The 9/11 World Trade Center (WTC) attack exposed thousands of people to toxic dust; this exposure has been associated with many illnesses, many of which have a long latency period and remain undetected until reaching an advanced stage with symptoms that affect one’s quality of life (1-4). A major component of the toxic dust exposure was asbestos, which was present in the fireproofing material used in the WTC (5,6). Asbestos exposure is associated with respiratory diseases such as lung cancer and asbestosis (7) and increased risk of cardiovascular disease (8).

The WTC dust exposure affects many different organ systems; two of the most prominent are the respiratory and cardiovascular systems (1,4,9-11). Studies have shown that firefighters with WTC exposure had higher age-adjusted incident rates of cardiovascular disease (1) and reduction in pulmonary function (10), with visible lung abnormalities on chest CT (4) and lower airway disease (12). A variety of abnormalities of the lung have been seen in firefighters with WTC exposure, including emphysema (5.9% of participants), pleural thickening (3.0%), and to a lesser extent pulmonary fibrosis (0.6%) (4). A more recent study of WTC responders found pleural abnormalities in 21% of the cohort (13). There was an increased risk of adverse cardiovascular events in firefighters with WTC exposure (1), although another study of police officers with WTC exposure found no increase of CAC after 5 years (14). CAC and aortic valve calcification (AVC) have both been shown to be independent prognostic risk factors of cardiovascular events and mortality (15) and can be measured on LDCT scans (16,17). A key question of WTC-related research is whether the amount of WTC dust exposure is related to the development and progression of disease. Previous WTC-related studies have found preliminary relationships between the amount of WTC exposure and various diseases. A study of firefighters found a linear relationship between the arrival time at the WTC site and decline in lung function measured by spirometry (10), and a later study found that high-intensity WTC exposure is associated with increased risks of air trapping, emphysema, and bronchial wall thickening (4). Another study on the WTC GRC found no association between WTC exposure duration or early arrival time at the WTC with pleural abnormalities on CT scans on a sub-cohort of 1453 members of the WTC GRC, from 2003-2012 (13), but another study found that reduced lung function from spirometry was lower in those who arrived early at the WTC and was associated with proximal airway disease (18). A recent study of firefighters showed an association between arrival time and duration at the WTC site with risk of cardiovascular disease (1). Male workers who arrived on the WTC site on 9/11 had a higher risk of heart disease compared to those who arrived after 9/18 (19), but this was not significant. A later study using a dust exposure questionnaire reported similar results (20). A study of workers in Italy exposed to asbestos reported an increase in all-cause mortality, and specifically lung and pleural cancers (21).

Many respiratory and cardiovascular diseases can be identified and quantified on CT and low-dose chest CT (LDCT) scans used for lung cancer screening. Emphysema has been quantified from CT scans by computing the percentage of lung volume below a set threshold (22), typically -910 HU. ILD has been automatically quantified by analyzing the texture of the lung parenchyma (23,24); one algorithm, CALIPER, used texture analysis of volumes of interest in the lung to classify regions into one of five different characteristic CT patterns (25). There are preliminary automated tools to quantify pleural thickening (26-28). Automated software for measuring CAC from chest CT has also been developed and showed good agreement with the current reference standard, Agatston scoring (29,30).

CT scans provide an opportunity to identify and evaluate the presence of early disease prior to the onset of symptoms in the WTC general responders’ cohort (GRC), as well as progression when multiple scans are obtained over time. Radiologists can visually and sometimes quantitatively assess diseases on CT scans, but these often have high variability, are time consuming to perform, or both (31-35). With the increase in computing power and advances in image analysis and machine learning, automated tools have been developed to assess a wide variety of diseases on CT scans (36-40).

In this work, we applied automated methods to quantify two respiratory diseases, emphysema ad interstitial lung disease, and two cardiovascular conditions, hypertension and coronary artery calcifications, in a cohort of WTC responders and a matched cohort of lung screening participants, to assess whether WTC exposure impacted the incidence rate or progression of these diseases.

## Methods

### Data

This study was approved by our institutional review board (IRB) and was HIPAA-compliant. We obtained a list of members of the WTC General Responders Cohort (GRC) from the WTC Data Center who received care at our institution through 2021. We queried our institution’s imaging archive for CT scans meeting the following criteria: 1) a patient on the list of WTC GRC members, 2) a CT scan that was non-contrast of the chest, and 3) slice thickness of 2.5 mm or less. The requirement for non-contrast CT scans with a slice thickness of 2.5 mm or less was to meet the image quality requirements of the automated software tools.

We curated a dataset of participants enrolled in the lung screening program at our institution that were matched on age, smoking status, follow-up time, and whether the participant had one or more CT scans to members of the WTC GRC for whom we had CT scans meeting the criteria above.

### Automated analysis

For pulmonary analyses, we used the open-source Chest Imaging Platform (CIP)^1^, an evolution of Airway Inspector. This software was developed primarily for quantitative CT analysis of lung disease, particularly chronic obstructive pulmonary disease (COPD). This is available as a docker container and ran on a workstation with an AMD Ryzen Threadripper PRO 5955WX with 128 GB of RAM. For cardiac analyses, we used research software developed by Cornell University (41,42) which we ran on a computer with an Intel Xeon Gold 6134 with 384gb of RAM. Images were converted from DICOM to the required image format for both software tools.

### Statistical analyses

Continuous variables were summarized using median and interquartile range(IQR) and compared across groups using Kruskal-Wallis test. Categorical variables were summarized by frequency and percentages and compared using chi-squared test or Fisher’s exact test as appropriate. All measures were assessed for normality and transformations were applied where necessary.

Controls were selected for each case using a systematic matching algorithm to ensure comparability. Cases were matched to controls based on sex, smoking status, visit type (one vs. two or more), and within 5 years difference of age. In addition, controls were required to have a follow-up time at least as long as that of the corresponding case. In instances where multiple controls met the matching criteria, one was randomly selected from the eligible pool to minimize selection bias. The matching process was conducted iteratively without replacement.

A linear mixed-effects model was employed to assess whether longitudinal changes in automated pulmonary and cardiovascular measurements derived from repeated CT scans differed between the WTC GRC and lung screening groups. Follow-up time was defined as time since initial CT scan with adjustment for race and current smoking status. Random intercept and slope were used to account for within-subject correlation across repeated measures and between-subject heterogeneity in the rate of change over time. Using these models, we calculated the estimated marginal means of outcome measures for comparison between WTC GRC and lung screening groups. Interaction terms between time and group (WTC GRC vs lung screening) were explored to assess potential differences in the trajectory of these measures across groups. Separate models were fitted for each pulmonary (emphysema LAA950; interstitial lung disease HAA250) and cardiovascular outcome (coronary artery calcification Agatston score; pulmonary hypertension PA ratio).

A two-sided p-value of < 0.05 was used to indicate statistical significance. All analyses were conducted using SAS (v9.4, SAS Institute, Cary, NC) and R (v4.3.0, R Foundation for Statistical Computing, Vienna, Austria).

## Results

There was a total of 4909 chest CT scans of members of the WTC GRC meeting our criteria. Of these 4909 chest CT scans, the emphysema and ILD analysis successfully completed on 4370 (89.0%) scans, while the coronary artery calcification was successful on 2122 (43.2%) scans, and both analyses were successful on 2023 scans (41.2%). There were 3855 members of the GRC for which we could obtain both chest CT images and clinical data. Of these, there were 2284 members for which we could obtain pulmonary disease measurements on at least one scan, 1246 members for which we could calculate cardiac measurements, and 1220 members for which we could successfully compute all four disease measures on at least one scan. Table 1 lists the longitudinal scans available for each GRC member; Table 1a lists those members for which we were able to compute pulmonary measurements, and Table 1b lists those members for which we were able to compute cardiac measurements. Although most members had a single CT scan, there were a substantial number with two or more scans available for longitudinal analysis (1070 members with pulmonary measurements and 724 members with cardiac measurements).

**Table 1.** Breakdown of number of CT scans available for each WTC GRC member.

*(a) Among 2284 members with at least one scan with successful pulmonary measurements*
| **Num CT scans** | **Num GRC members** | **Percent (%)** |
| --- | --- | --- |
| **1** | 1214 | 53.2 |
| **2** | 520 | 22.8 |
| **3** | 279 | 12.2 |
| **4** | 135 | 5.9 |
| **5** | 66 | 2.9 |
| **6 or more** | 70 | 3.1 |
| **Total** | 2284 |  |

*(b) Among 1246 members with at least one scan with successful cardiac measurements*

| **Num CT scans** | **Num GRC members** | **Percent (%)** |
| --- | --- | --- |
| **1** | 522 | 41.9 |
| **2** | 302 | 24.2 |
| **3** | 194 | 15.6 |
| **4** | 105 | 8.4 |
| **5** | 56 | 4.5 |
| **6 or more** | 67 | 5.4 |
| **Total** | 1246 |  |

We successfully identified eligible controls for 557 cases based on the pre-specified matching criteria. The demographic characteristics of the matched case-control groups are shown in Table 2. Of the 557 matched pairs, 457 (82%) of them were male and 18% female. The median age was 54 years, with an interquartile range of 48 to 60 years for the WTC GRC group and 49 to 60 years for the control lung screening group. Although matching was performed based on smoking history (whether individuals had ever smoked), current smoking status differed significantly between the groups (P < .0001). In the WTC GRC group, 23% were current smokers, 30% were former smokers, and 47% were never smokers. In contrast, 34% of the control lung screening group were current smokers, 19% were former smokers, and 47% were never smokers.

**Table 2.** Demographics 557 members of WTC GRC and 557 members of the control lung screening cohort.

| Demographic characteristics | WTC GRC | Control Lung Screening | P-value |
| --- | --- | --- | --- |
| N | 557 | 557 |  |
| Sex |  |  |  |
| Male | 457 (82%) | 457 (82%) | 1.00 |
| Female | 100 (18%) | 100 (18%) |  |
| Age*, y | 54 (48, 60) | 54 (49, 60) |  |
| Smoking |  |  |  |
| Current smoker | 126 (23%) | 189 (34%) | <.0001 |
| Former smoker | 169 (30%) | 106 (19%) |  |
| Never smoker | 262 (47%) | 262 (47%) |  |
| Race |  |  |  |
| American Indian/Alaska Native | 1 (0%) | 3 (1%) | <.0001 |
| Asian | 6 (1%) | 74 (13%) |  |
| Native Hawaiian or Other Pacific Islander | 0 (0%) | 0 (0%) |  |
| Black or African America | 50 (9%) | 107 (19%) |  |
| White | 339 (61%) | 308 (55%) |  |
| More than One Race | 124 (22%) | 65 (12%) |  |
| Unknown or Not Reported | 37 (7%) | 0 (0%) |  |
| Arrival Time |  |  |  |
| 9/11 Dust Cloud | 94 (17%) |  |  |
| 9/11 No Dust Cloud | 62 (11%) |  |  |
| 9/12 & 9/13 | 168 (30%) |  |  |
| 9/14 - End of Sept | 167 (30%) |  |  |
| Oct and beyond | 50 (9%) |  |  |
| Unknown | 16 (3%) |  |  |
| Average length of follow-up (SD), year | 3.61 (4.07) | 1.79 (3.01) | <.0001 |

Race distributions also varied significantly between the two groups (P < .0001). The WTC GRC group had a higher proportion of White participants (61%) compared to 55% in the control lung screening group. A higher proportion of Black or African American participants was observed in the control lung screening group (19%) compared to the WTC GRC group (9%). There were also notable differences in the representation of Asian participants (1% in WTC GRC vs. 13% in control). Among the 557 WTC GRC, arrival time for the WTC GRC group was recorded across various time points, with 17% exposed to the 9/11 Dust Cloud, 11% arriving after the dust cloud on 9/11, 30% arriving on 9/12 or 9/13, 30% arriving between 9/14 and the end of September, and 9% arriving in October or later.

### Extent of disease on initial CT scan

Table 3 summarizes pulmonary and cardiac measures from the first CT scan for both the WTC GRC and control lung screening groups. Among the 184 participants with available cardiac findings, the median Agatston score, a measure of coronary artery calcification, was significantly higher on the first CT scan in the WTC GRC group (median: 68.70, IQR: 18.31, 258.92) compared to the control group (median: 8.65, IQR: 0.12, 120.04) (p<0.001). PA ratio, a measure of pulmonary hypertension, was similar between the WTC GRC group (median: 0.79, IQR: 0.73, 0.88) and the control group (median: 0.79, IQR: 0.72, 0.86) (p=0.25).

**Table 3.** Summary of cardiovascular and respiratory disease measurements on the WTC GRC members on the first available CT scan.

| Measures on first CT scan | WTC GRC | Non-WTC GRC | P-value |
| --- | --- | --- | --- |
| **With available cardiac findings** | 184 |  |  |
| **Coronary Artery Calcification** |  |  |  |
| Agatston score* | 68.70 (18.31, 258.92) | 8.65 (0.12, 120.04) | <0.001 |
| **Pulmonary Hypertension** |  |  |  |
| PA ratio* | 0.79 (0.73, 0.88) | 0.79 (0.72, 0.86) | 0.249 |
| **With available pulmonary findings** | 556 |  |  |
| **Emphysema** |  |  |  |
| LAA950* | 0.09 (0.01, 0.19) | 0.03 (0.01, 0.09) | <0.001 |
| **Interstitial lung diseases** |  |  |  |
| HAA250* | 0.02 (0.02, 0.03) | 0.02 (0.01, 0.02) | <0.001 |

\*Median (IQR)

Among the 556 participants with available pulmonary findings, significant differences were observed for emphysema and interstitial lung diseases. The LAA950 score, which reflects the extent of emphysema, was significantly higher in the WTC GRC group (median: 0.09, IQR: 0.01, 0.19) compared to the control group (median: 0.03, IQR: 0.01, 0.09) (p<0.001), suggesting more extensive emphysema in the WTC GRC group. Similarly, the HAA250 score, a measure of interstitial lung disease, was also significantly higher in the WTC GRC group (median: 0.02, IQR: 0.02, 0.03) compared to the control group (median: 0.02, IQR: 0.01, 0.02) (p<0.001), indicating more extensive ILD among WTC GRC.

Overall, these results indicate that participants in the WTC GRC group exhibit a higher burden of coronary artery calcification, emphysema, and interstitial lung disease compared to those in the control group, while no significant difference was observed for pulmonary hypertension.

### Progression of diseases

As shown in Table 4, at baseline, the average LAA950 was significantly higher in the WTC GRC group compared to the control lung screening group (0.12 vs. 0.075, P < 0.001), suggesting that WTC GRC had more extensive emphysema. Over time, the WTC GRC group showed a significant decrease in LAA950 score, with a yearly reduction of -0.011 (95% CI: -0.012, -0.009, P < 0.001). In contrast, the control group exhibited a slight but statistically significant increase in LAA950 over time, with a yearly increase of 0.0042 (95% CI: 0.0021, 0.0063, P < 0.001), indicating worsening of emphysema.

**Table 4.** Extent and progression of respiratory and cardiovascular disease measures for members of the WTC GRC and the control group.

| a) Emphysema (LAA950) |  |  |  |  |  |  |
| --- | --- | --- | --- | --- | --- | --- |
|  | At Baseline | 95% CI | p-value | Δ/year | 95% CI | p-value |
| WTC GRC | 0.12 | (0.11, 0.14) | <0.001 | -0.011 | (-0.012, -0.009) | <0.001 |
| Control lung screening | 0.075 | (0.06, 0.09) |  | 0.0042 | (0.0021, 0.0063) |  |

| b) Interstitial lung diseases (HAA250) |  |  |  |  |  |  |
| --- | --- | --- | --- | --- | --- | --- |
|  | At Baseline | 95% CI | p-value | Δ/year | 95% CI | p-value |
| WTC GRC | 0.027 | (0.025, 0.028) | <0.001 | -0.0006 | (-0.0007, -0.0005) | <0.001 |
| Control lung screening | 0.019 | (0.017, 0.021) |  | -0.0001 | (-0.0004, 0.0001) |  |

| c) Pulmonary Hypertension (PA ratio) |  |  |  |  |  |  |
| --- | --- | --- | --- | --- | --- | --- |
|  | At Baseline | 95% CI | p-value | Δ/year | 95% CI | p-value |
| WTC GRC | 0.77 | (0.72, 0.82) | 0.91 | -0.0002 | (-0.0047, 0.0042) | 0.85 |
| Control lung screening | 0.77 | (0.71, 0.83) |  | -0.0009 | (-0.0067, 0.0048) |  |

| d) Log CAC Agatston score |  |  |  |  |  |  |
| --- | --- | --- | --- | --- | --- | --- |
|  | At Baseline | 95% CI | p-value | Δ/year | 95% CI | p-value |
| WTC GRC | 4.47 | (3.47, 5.48) | 0.045 | 0.09 | (0.033, 0.14) | <0.001 |
| Control lung screening | 2.88 | (1.74, 4.03) |  | 0.26 | (0.19, 0.33) |  |

All models adjusted for race and current smoking status

At baseline, the HAA250 score was also significantly higher in the WTC GRC group compared to the control group (0.027 vs. 0.019, P < 0.001). Figure 1 depicts the progression of the pulmonary disease measures over time for the WTC GRC group and control group. The WTC GRC group demonstrated a significant annual decrease in HAA250 (−0.0006, 95% CI: -0.0007, -0.0005, P < 0.001), while the control group did not exhibit any statistically significant change in HAA250 over time (−0.0001, 95% CI: -0.0004, 0.0001).

**Figure 1.**
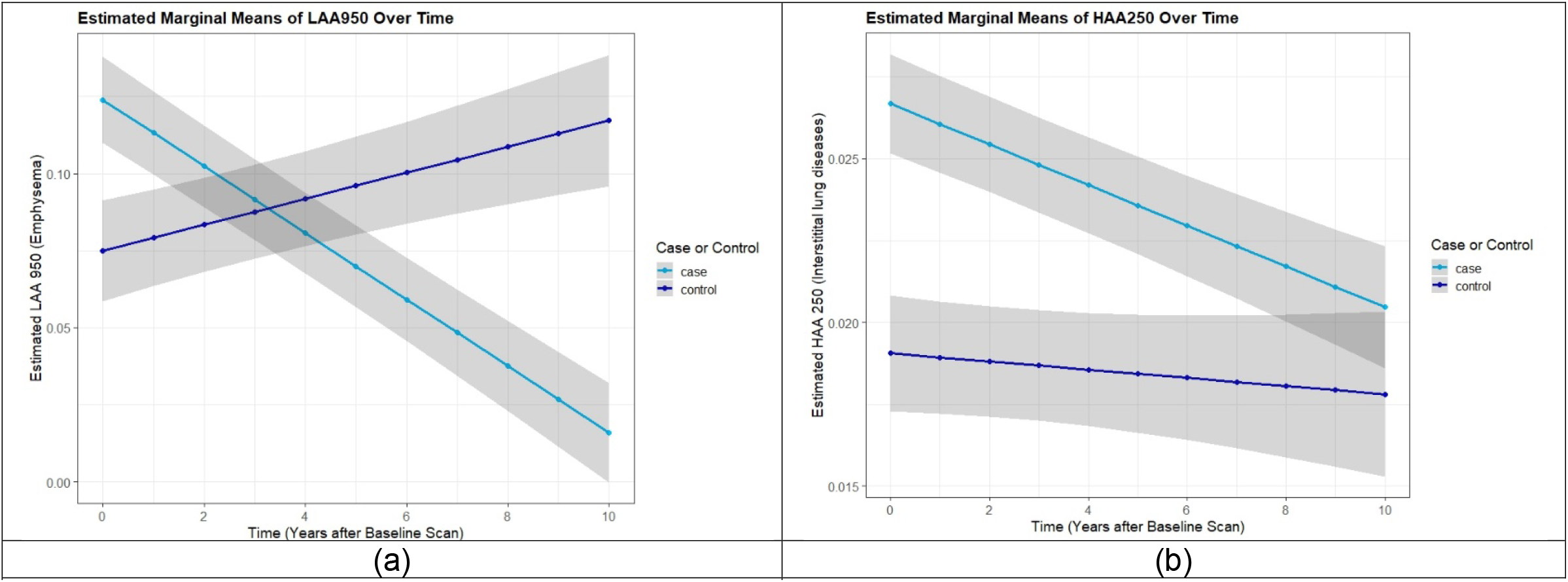
Respiratory measurements over time for the WTC GRC (Case) and lung screening control groups (control). a) Estimated mean emphysema score (LAA950) and b) Estimated mean interstitial lung disease measure (HAA250)

At baseline, the PA ratio was similar between the WTC GRC and control groups (p=0.91). No significant differences were observed in the progression of pulmonary hypertension (PA ratio) between the groups (P = 0.85).

Agatston score was log-transformed prior to analysis due to the skewed distribution. The baseline log-transformed CAC Agatston score was significantly higher in the WTC GRC group compared to the control group (4.47 vs. 2.88, P = 0.045), indicating more extensive coronary artery calcification. Both groups showed significant increases in the CAC score over time, with the WTC GRC group showing an annual rate of increase of 0.09 (95% CI: 0.033, 0.14) and the control group showing a greater increase of 0.26 (95% CI: 0.19, 0.33).

Figure 2 depicts the progression of the cardiac measures over time for the WTC GRC group and control group. The rate of progression was significantly different between the two groups (p<.001) with the control group showing faster progression.

**Figure 2.**
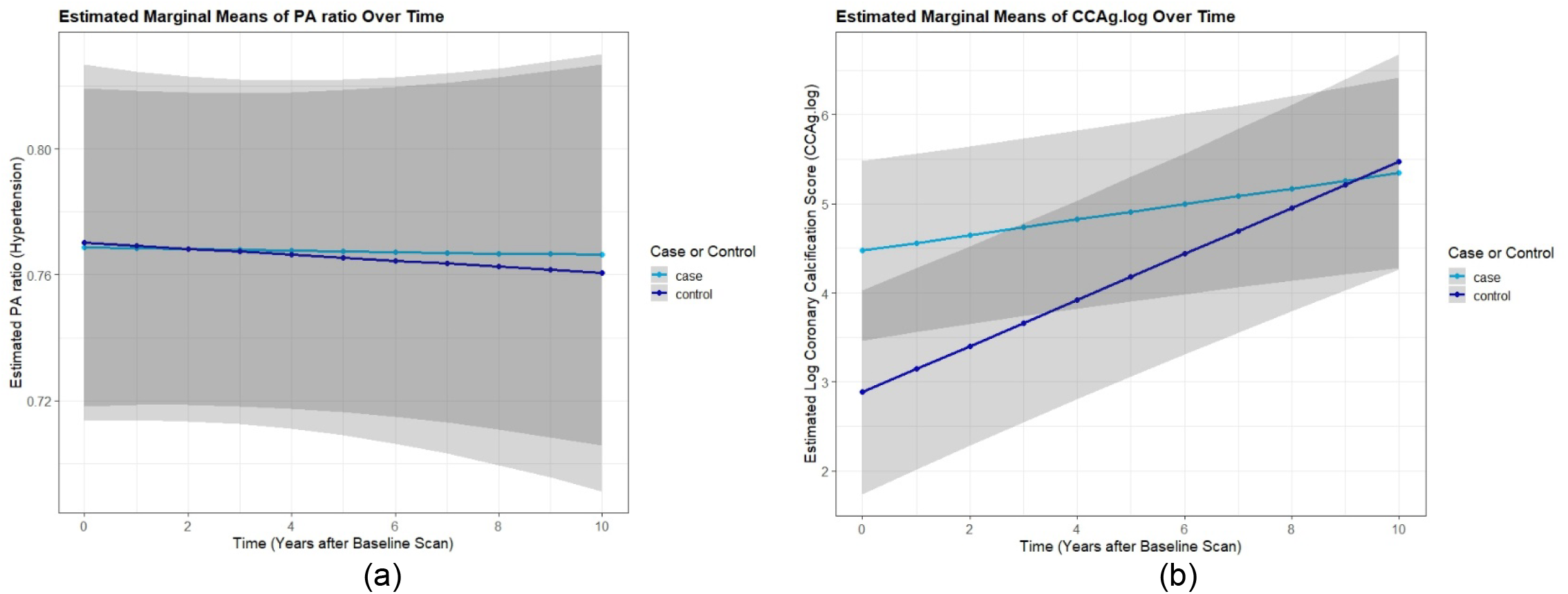
Cardiac measurements for the WTC GRC group (Case) and control lung screening group (control) over time. a) Estimated mean pulmonary artery to aorta ratio (PA ratio) and b) log-transformed Agaston score.

## Discussion

This analysis showed that members of the WTC GRC had elevated levels of coronary artery calcification, emphysema, and interstitial lung disease compared to the lung screening control group on the initial CT scan. The median time between 9/11 and the initial CT scan included in this study for the WTC GRC was about 10 years, which is a long enough time that any effect of WTC exposure on these diseases would already be seen. It also suggests that the burden of these diseases remains elevated even after 10 years; this supports the need for long-term health monitoring and sustained surveillance programs for WTC GRC members. A previous study by Weakley et al. showed that emphysema and COPD were more prevalent in the WTC-exposed fire fighters compared to the general population 5 years after 9/11, increasing or remaining stable through year 8 (43). While a previous study by Wanahita et al. did not find an increased prevalence of coronary artery disease in WTC-exposed police officers 5 years after 9/11, but the authors note that majority of the police officers were below 40 years of age (14); however another study on WTC-exposed fire fighters by Cohen et al. found an association between WTC exposure and long-term cardiovascular disease risk (1).

The results on the progression of disease were negative – although there was a statistically significant decrease in emphysema and interstitial lung disease, we know that these diseases are not reversible. This is likely attributable to differences in CT scanners and acquisition protocols, Chen-Mayer et al. found that there were differences in lung density measurements across different scanners and protocols, which could be reduced by applying standardization procedures (44). There was no significant difference observed between the WTC GRC group and the control group for the progression of hypertension which correlates with the denial of a petition to add hypertension as a WTC health condition (45). While both the WTC GRC group and control group had increasing coronary artery calcification (CAC) over time, surprisingly the control group showed faster progression. This could again to attributable to differences in CT scanners and protocols; the lung screening group tended to have more recent CT scans with thinner slice thickness, which may be more sensitive for detection of CAC. Lastly, the etiology of pulmonary diseases may differ between the two groups due to the nature of exposure, potentially result in different clinical manifestations and progression of disease, complicating the comparative analysis.

In computing the automated disease measures, we encountered algorithm failures in the coronary analysis in over half of the CT scans. The lower success rate of the coronary artery analysis compared to the emphysema and ILD analyses is due to the higher difficulty of finding small calcium deposits in non-gated CT images – normally coronary artery calcifications are measured on gated CT imaging to eliminate artifacts caused by heart motion, but for this analysis, we are using chest CT images obtained for other purposes. The specific algorithm also requires the segmentation of a number of anatomical structures in the chest, including the lungs, airways, and ribs, which are more difficult than just segmenting the lungs. Both software for the emphysema/ILD measures and CAC measures utilized model-based methods as opposed to deep learning methods. There have been many advancements in deep-learning based methods in the past few years and newer methods may provide a higher success rate.

## Data Availability

Data produced in this present study are not available, though the underlying clinical and image data are available upon request to the World Trade Center General Responders Data Center.

## Acknowledgements

This study was funded by an NIH/CDC grant R21OH012244.

## Footnotes

1 Available from https://chestimagingplatform.org/index.htm

